# The +57C>T substitution in microRNA-184 is associated with microphthalmia, retinal detachment, and altered ocular development

**DOI:** 10.64898/2026.06.25.26355554

**Authors:** Yuejia Huang, Yanjun Zhang, Sonya Zhang, Kadia Lissit, Audrey Talley-Rostov, Charles C. Lin, Patricia S. Tsai, Alice Hong, Anshuman Agrawal, Jonathan Thomas, Lo-Yu Chang, Michael Sulewski, Luisa Cochella, Jinchong Xu, Allen O. Eghrari

**Author notes:** Equal contributors (Jinchong Xu, Allen O. Eghrari). **Corresponding Authors:** Allen O. Eghrari, MD, MPH, Department of Ophthalmology, Johns Hopkins Univ. School of Medicine, 1800 Orleans Street, Wilmer 120, Baltimore, MD, 21287, USA,; Jinchong Xu, MD, PhD, Department of Neurology, Johns Hopkins Univ. School of Medicine, 733 N. Broadway, Miller Research Building 769, Baltimore, MD, 21205-1832, USA. **Address for Reprints**: Allen O. Eghrari, MD, MPH, Department of Ophthalmology, Johns Hopkins University School of Medicine, 1800 Orleans Street, Wilmer 120, Baltimore, MD 21287, USA. **Commercial Relationships Disclosure:** Y. Huang, Revivify Innovations (O); Y. Zhang, None; K. Lissit, None; S. Zhang, None; A. Talley-Rostov, None; C.C. Lin, None; P.S. Tsai, None; A. Hong, None; A. Agrawal, None; J. Thomas, None; L. Chang, None; M. Sulewski, Revivify Innovations (O); L. Cochella, None; J. Xu, Revivify Innovations (O); A.O. Eghrari, Revivify Innovations (O).

## Abstract

**Purpose:** To expand the clinical and mechanistic understanding of the +57C>T seed-region mutation in miR-184 causing EDICT (endothelial dystrophy, iris hypoplasia, congenital cataract, and stromal thinning) syndrome.

**Design:** Cross-sectional analysis and laboratory confirmation

**Participants:** 18 members of a four-generation family with known +57C>T miR-184 status

**Methods:** We used optical biometry, corneal topography, and medical history to characterize the clinical phenotype. Carrier effects on ocular biometric measurements were estimated using polygenic linear mixed models incorporating a pedigree-derived kinship matrix, adjusted for age and sex. Patient-derived and control induced pluripotent stem cells (iPSCs) were generated and differentiated into corneal endothelial cells (CECs).

**Main Outcome Measures:** Axial length, keratometry (in diopters), white-to-white corneal diameter, topography mapping, central and peripheral corneal thickness, and history of retinal detachment or corneal transplant were compared between mutation carriers and noncarriers, adjusting for age and sex. Cellular analysis was conducted with immunostaining (ZO-1, ATP1A1), morphometric quantification, qRT-PCR of endothelial markers, and transendothelial electrical resistance (TEER).

**Results:** 10 of 18 family members were heterozygous for +57C>T, with retinal detachment occurring in 5/10 affected individuals compared to 0/8 unaffected individuals (p=0.04). Affected eyes had 2.2 mm shorter axial length (p=0.02), 9.3 D steeper mean keratometry (p=0.004), 1.6 mm smaller horizontal corneal diameter (p=0.0001), and 139-µm thinner central corneas (p=0.003). Mutant iPSC-derived CECs were associated with irregular borders, increased cell and nucleus area, widened intercellular gaps, disrupted ATP1A1 membrane localization, and reduced barrier function on TEER (all p<0.05). Gene expression analysis showed downregulation of COL4A1, COL4A3, and AQP1 with upregulation of COL8A1.

**Conclusions:** The miR-184 +57C>T mutation produces a broad ocular phenotype that includes smaller, thinner corneas and microphthalmia. Mechanistically, it disrupts CEC junctional integrity, extracellular matrix and pump-related genes, supporting a role for miR-184 in coordinated anterior-posterior eye morphogenesis.

## Introduction

Endothelial dystrophy, iris hypoplasia, congenital cataract and stromal thinning syndrome (EDICT syndrome, OMIM #614303) is a rare, autosomal dominant condition^1,2^ resulting from a +57C>T substitution in the seed region of miR-184.^3^ Its prevalence is unknown, but families have been identified with this mutation in both the United States and Europe.^1,4,5^ Despite features that overlap with other autosomal dominant anterior segment syndromes, EDICT syndrome is uniquely characterized by the simultaneous presence of endothelial dystrophy, iris hypoplasia, congenital cataracts, and corneal stromal thinning.^2^ The constellation of symptoms was initially localized to a gene on chromosome 15q22.1-25.3 between markers D15S993 and D15S202, and later associated with a causal variant, a single base substitution +57C>T in the seed region of miR-184. ^2,3^ This was the first ocular condition to be associated with a mutation in a microRNA.

Since the initial description of EDICT syndrome, the emergence of retinal detachment among several family members has raised the question of whether the phenotype extends beyond the anterior segment.^3^ We sought to more broadly characterize the phenotype of this condition at both the cellular and organ (eye) levels.

Prior zebrafish studies show that loss of miR-184 leads to microphthalmia, cataract, and altered crystallin homeostasis, in part via upregulation of the cell-cycle inhibitor p21, which causes impaired cellular proliferation and lens fibrosis.^6^ It is possible, therefore, that mutant miR-184 disrupts the developmental gene expression cascades critical to the morphogenesis of the anterior and posterior segments. Therefore, we hypothesized that eyes affected by EDICT syndrome experience altered phenotypes throughout the lifespan, with functional changes at both a cellular and organ level.

Notably, recent transcriptome-wide profiling in human corneal endothelial cells carrying miR-184 variants has further supported a primary endothelial disease mechanism, identifying broad dysregulation of pathways relevant to endothelial identity and homeostasis.^7^ These findings provide complementary molecular context for the cellular phenotypes we evaluate here using iPSC-derived corneal endothelial cells (CECs).

## Methods

### Participants

A large family with inherited EDICT syndrome in an autosomal dominant pattern of inheritance across four generations (Figure 1A) were included. Participants had previously confirmed affection status through genetic testing including Sanger sequencing.^2^ In this study, “affected” refers to individuals with the +57C>T substitution in the seed region of miR-184, whereas “unaffected” refers to individuals tested to be negative for the genetic variant. There is 100% penetrance in this family, such that “affected” and “unaffected” refer to both the genotype and phenotype. Sex information was self-reported.

**Figure 1.**
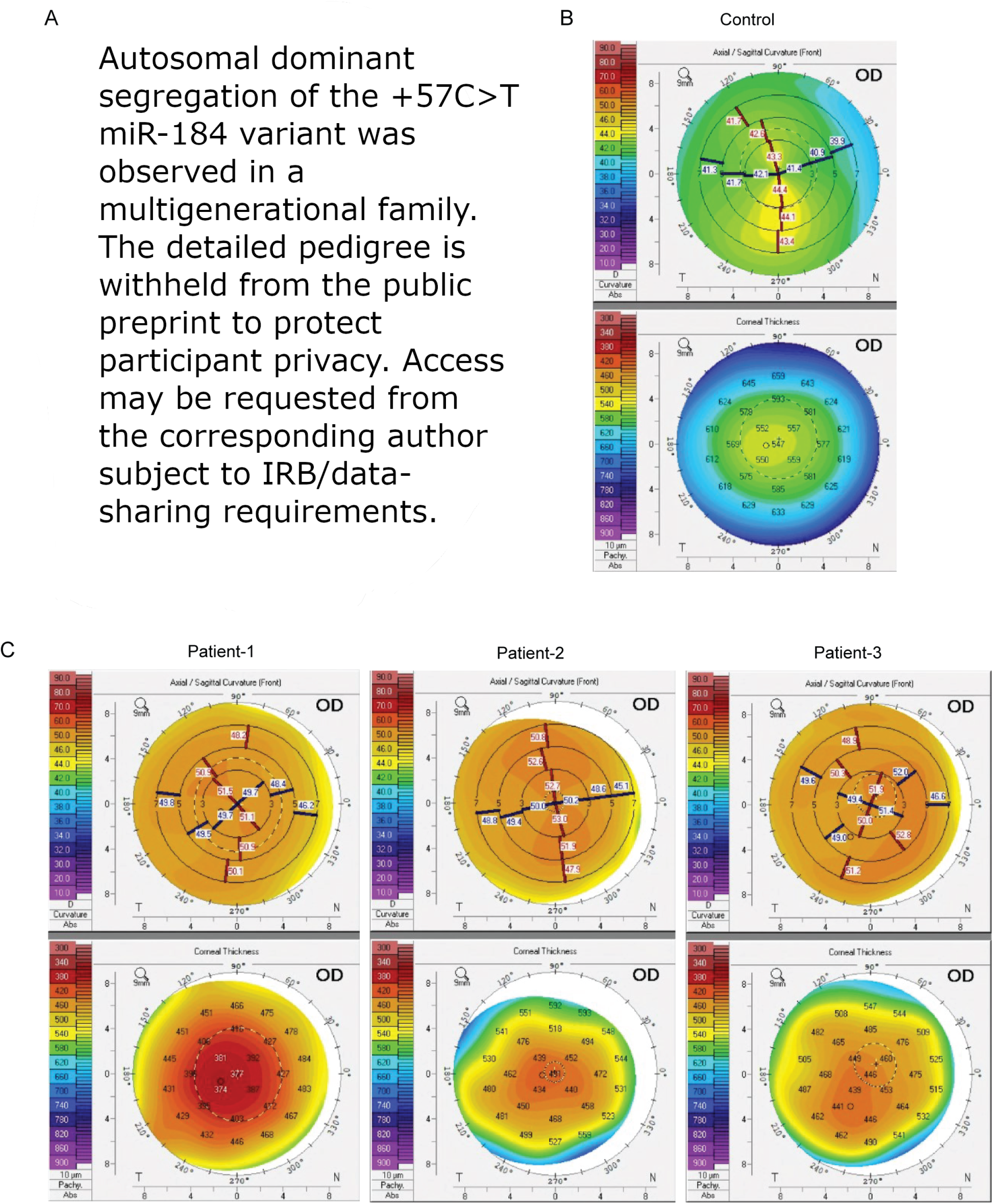
The +57C>T substitution in the seed region of miR-184 disrupts corneal structure and function and alters ocular development. **(A)** Pedigree of family affected by +57C>T substitution in the seed region of miR-184. **(B, C)** Corneal topography with Scheimpflug imaging of the control vs patient corneas. Abbreviations: microRNA-184 = micro-ribonucleic acid 184.

13 of 18 family members provided corneal imaging data including optical biometry with IOL Master (Carl Zeiss Meditec, Inc., Dublin, CA, USA) or Lenstar (Haag-Streit USA, Mason, OH, USA). Corneal topography was conducted with Pentacam (OCULUS Inc., Arlington, WA, USA).

Induced pluripotent stem cells (iPSCs) were produced using a 10 mL blood sample from an affected family member aged 76–80 years. Institutional Review Board approval was obtained from the Johns Hopkins University School of Medicine. All study participants consented to the approved study protocol, which was conducted in accordance with the Declaration of Helsinki. Written informed consent was obtained from all individuals prior to enrollment.

### Cell lines

Peripheral blood mononuclear cells (PBMCs) were reprogrammed using a Sendai virus delivery system kit, according to the manufacturer’s instructions (Cytotune 2.0, Life Technologies, Carlsbad, CA, USA). iPSC colonies were cultured under feeder-free conditions on Matrigel-coated plates (Corning Life Sciences, Tewksbury, MA, USA) and maintained in mTeSR1 media (STEMCELL Technologies Inc., Vancouver, BC, Canada). iPSC lines and biological replicates. One patient-derived iPSC line harboring the +57C>T miR-184 mutation (from aan affected family member aged 76–80 years) and three unrelated healthy control iPSC lines (JHiPS-30516, JHiPS-70226, JHiPS-70351) were used in this study. Unless otherwise specified, experiments were performed using 3 independent differentiation batches per line (biological replicates). All experiments involving human iPSCs were conducted in accordance with JHUSOM policy, which requires that any research involving human iPSCs is subject to oversight by the JHU Institutional Stem Cell Research Oversight (ISCRO) Committee.

### Comparison of miR-184 expression in iPSCs, scleral, and corneal tissues

To confirm relative miR-184 expression between corneal endothelium and sclera, we obtained one unused DMEK graft (corneal endothelium with Descemet’s membrane) and an accompanying scleral rim from a single deidentified research-consented donor cornea. Pre-differentiated iPSC and iPSC-CEC samples (14 days post differentiation) from a control sample were also collected. Total RNA was extracted from each tissue using TRIzol reagent (Invitrogen, Carlsbad, CA, USA) following the manufacturer’s chloroform extraction protocol to preserve small RNA species. Mature hsa-miR-184 was quantified using the TaqMan MicroRNA Assay (Applied Biosystems, Foster City, CA, USA) on the CFX Connect Real-Time PCR Detection System (Bio-Rad, Hercules, CA, USA) with RNU6A (U6) snRNA as the endogenous small-RNA control. Relative expression was calculated by the ΔΔCt method, with pre-differentiated iPSC serving as the normalizing sample (set to 1.0). Reactions were run in technical triplicates.

### Northern blot detection of miR-184

Total RNA was extracted from pre-differentiated iPSC and iPSC-CEC (14 days post differentiation) samples using TRIzol reagent (Invitrogen, Carlsbad, CA, USA) following the manufacturer’s chloroform extraction protocol to preserve small RNA species. Northern blots were performed as previously described^8^ with minor modifications. Briefly, probes were synthesized as ssDNA oligos and 5′-^32^P-radiolabeled with T4 polynucleotide-kinase (New England Biolabs, Ipswich, MA, USA) and γ-^32^P-ATP (6000 Ci/mmol, Revvity, Waltham, MA). Unincorporated nucleotides were removed by G25 column purification (Cytiva, Marlborough, MA, USA). After semidry transfer (BioRad, Hercules, CA, USA) onto Hybond NX membranes (Cytiva), RNA was UV crosslinked three times with 120 mJ/cm² each, followed by chemical crosslinking using methylimidazole/EDC. Membranes were prehybridized with Church buffer (1 mM EDTA, 0.5 M Na_2_HPO_4_/NaH_2_PO_4_, 7% SDS) for 1 h at 65 °C in a hybridization oven, after which 25 pmol radioactively labeled probes were added. The temperature was lowered to 37 °C, and membranes incubated overnight. Membranes were washed at 37 °C three times with 1x SSC + 0.1% SDS for 10 min before exposure to a storage phosphor screen (Cytiva). All imaging was performed with a Typhoon phosphorimager (Cytiva).

### Rationale for CEC model

We focused mechanistic experiments on iPSC-derived corneal endothelial cells (CECs) because corneal endothelial dystrophy and barrier dysfunction are consistent clinical features in +57C>T miR-184–associated EDICT syndrome, and the corneal endothelium is a tractable human cell type for quantifying junctional integrity, extracellular matrix–related programs, and pump-associated membrane organization. This model was designed to test cell-autonomous consequences of the miR-184 seed mutation on endothelial morphology and function.

### Differentiation of iPSCs to Corneal Endothelial Cells

Differentiation was conducted by adapting a protocol developed by Ali and colleagues.^9^ For each iPSC line, CEC differentiation was performed three independent times (biological replicates), with each differentiation generating multiple wells/fields for downstream assays as described below. iPSCs were seeded on 35-mm Matrigel-coated plates. The iPSCs were grown for 2 days in medium (mTeSR1; STEMCELL Technologies, Inc.). On day 1, mTeSR1 media was replaced with dual Smad inhibitors media containing 50 ng/mL human recombinant Noggin (R&D Systems, Minneapolis, MN, USA), 10 µM SB431542 (MilliporeSigma, Burlington, MA, USA) and 1 µM Dorsomorphin in a CEC basal media of 80% DMEM-F12 (Life Technologies), 20% KSR (Life Technologies), 1% nonessential amino acids (Life Technologies), 1 mM L-glutamine (STEMCELL Technologies, Inc.), 0.1 mM β-mercaptoethanol (MilliporeSigma), and 8 ng/mL bFGF (MilliporeSigma). On day 3, dual Smad inhibitors media was replaced by cornea medium containing 0.1% B27 supplement (Life Technologies), 10 ng/mL recombinant human platelet derived growth factor-BB (PDGF-BB; PeproTech, Rocky Hill, NJ, USA), and 10 ng/mL recombinant human Dickkopf related protein-2 (DKK-2; R&D Systems) in the CEC basal media. On day 4 or later, the differentiating CECs were transferred to new plates and were grown in cornea medium for 10 additional days.

### Immunostaining of Corneal Endothelial Cells and Quantification

Cells were fixed in precooled methanol for 10 min. After three PBS washes, samples were incubated for 30 min at room temperature in 5% normal donkey serum. Next, samples were incubated overnight at 4 °C with the indicated primary antibodies of the tight junction protein-1 (ZO1), Na, K-ATPase alpha-1 subunit (ATP1A1) and then washed three times with PBST (0.2% Triton X-100 in PBS). The cells were then incubated for 1 h in the indicated secondary antibodies and washed three times. Samples were mounted on dishes with anti-fading mounting medium, and slides were inspected by microscope (LSM880; Carl Zeiss Inc., Thornwood, NY, USA). Immunostaining experiments were performed using CECs from three independent differentiations per line. For each condition, images were acquired from three wells per differentiation, with nine to twelve non-overlapping fields per well.

The following definitions were used when inspecting the slides:

- **Cell Area**: Surface area of a single corneal endothelial cell as observed under ZO-1 staining.
- **Nucleus Area**: Nucleus size of a single corneal endothelial cell as observed under DAPI.
- **Gap Distance**: Straight-line distance between adjacent cells.
- **Gap Area**: Extracellular space between cells, measured as the area outside the cytosol separating one cell from another.
- **Number of Patches**: Degree of clustering which means fewer patches suggest cell has lower continuity.
- **Patch Density**: Number of patches per unit area of the image. Lower patch density indicates fewer or more sparsely distributed patches, which means higher continuity.
- **Number of Peaks**: A larger number of peaks suggests greater structural or intensity heterogeneity within the cell. In contrast, a more evenly distributed structure or intensity profile will result in fewer peaks. Cells with a smooth or homogeneous structure typically produce a lower number of peaks, while cells with irregular structures, clusters, or localized intensity variations show a higher peak count.
- **Average Peak Intensity**: The average peak intensity represents the mean brightness or intensity of all detected peaks in the radial intensity profile. This metric provides a measure of the signal strength at the detected regions of interest (peaks). Higher average peak intensity might indicate stronger fluorescence or more pronounced features in the cell’s structure.

### Quantitative Real-Time Polymerase Chain Reaction (qRT-PCR)

The total RNA was extracted using reagent (TRIzol; Invitrogen, Carlsbad, CA, USA) according to the manufacturer’s instructions. The RNA was quantitated on a spectrophotometer (NanoDrop Lite; Thermo Fisher Scientific, Wilmington, DE, USA). First-strand cDNA was synthesized using a kit (Superscript III; Invitrogen, Thermo Fisher Scientific, Carlsbad, CA, USA) according to the manufacturer’s instructions.

qRT-PCR was performed on the QS6 Real-Time PCR System (Applied Biosystems, Foster City, CA, USA). The expression of CEC markers was quantitated using Power SYBR Green PCR Master Mix (Life Technologies). GAPDH was used as the endogenous control. The delta-delta Ct method was used to determine the relative expression, normalized against GAPDH, as reported previously. All the primers used in the qRT-PCR analysis were designed using a real-time PCR tool (Integrated DNA Technologies, Coralville, IA, USA) and are available upon request.

qRT-PCR was performed using RNA from three independent differentiations per line. Relative expression was calculated by ΔΔCt and summarized as mean ± SEM across biological replicates.

### Transendothelial Electrical Resistance (TEER)

CECs were differentiated as described. After 7 days, 150,000 cells were seeded in 24-well plates (6.5-mm diameter, 0.4-µm pore; Corning Life Sciences), previously coated with Matrigel. TEER was measured with an EVOM2 epithelial volt-ohmmeter (World Precision Instruments, Sarasota, FL, USA) for 30 days or until readings reached a steady state. TEER measurements were normalized to the value of the control wells (growth media only). The reported values represent the average reading of at least three independent differentiation batches.

### Statistical analysis

Descriptive analyses and multivariable regression analyses, controlling for age and sex, were conducted to characterize the disease phenotype. To verify that within-family correlation did not influence our findings, each multivariable regression was fit as a polygenic linear mixed model incorporating a kinship matrix derived from the family pedigree, using the kinship2 and coxme packages in R.^10^ As a further sensitivity analysis, each model was re-fit on a subset restricted to one individual per sibship. The measured value was averaged across both eyes for measures of axial length, white-to-white, corneal thickness, and corneal curvature. Eyes with history of corneal transplant were excluded from corneal analyses, and eyes with known history of scleral buckle were excluded from axial length analyses. Ocular biometric analyses were performed as complete-case analyses using available imaging data. Corneal imaging data were available for 13 of 18 family members, including 9 affected and 4 unaffected individuals; the remaining 5 family members without imaging data were not imputed.

For cell-based assays, the primary unit of analysis was the independent differentiation (biological replicate), with technical replicates (wells/fields/cells) averaged within each differentiation prior to statistical testing. Findings should be carefully interpreted with the understanding that multiple-comparison adjustment was not performed, given the modest sample size and the need to balance the risk of Type I and Type II errors.

## Results

In this multi-generational family comprising 18 living individuals (Figure 1A), 10 (56%) were heterozygous for the +57C>T miR-184 mutation.

### Clinical Phenotype

Among affected family members, 5 of 10 (50%) reported undergoing surgery for retinal detachment in at least one eye, compared to 0 of 8 (0%) unaffected family members (p=0.04, Fisher’s exact test).

A total of 9 affected and 4 unaffected family members underwent optical biometry and/or corneal topography (Table 1). Linear mixed models adjusted for age, sex, and pedigree-derived relatedness revealed that eyes harboring the mutation had 2.2 mm shorter axial length (95% CI, 0.40 to 4.03 mm; p=0.022) and a 1.6 mm reduction in corneal diameter (95% CI, 1.10 to 2.11 mm; p<0.001), as measured by white-to-white. Scheimpflug corneal topography demonstrated 9.3 D increased corneal curvature (95% CI, 3.83 to 14.78 D; p=0.004) and a 139-µm reduction in central corneal thickness (95% CI, 66.8 to 210.6 µm; p=0.003), both also after adjustment for age, sex, and relatedness. Notably, compared to unaffected family members (Figure 1B), the pattern of corneal curvature on corneal topography in affected members is not only one of central thinning, but also peripheral thinning, with regular astigmatism (Figure 1C). Additional topography images are included in Supplemental Figure S1.

**Table 1.** Carrier-effect estimates associated with the +57C>T substitution in the seed region of miR-184 (n = 13 measured individuals from one four-generation family). Group descriptive statistics are reported as mean ± SD of per-individual values, with each individual’s value calculated as the average across both eyes. Affected refers to individuals heterozygous for the +57C>T miR-184 variant; unaffected refers to family members negative for the variant. *Carrier effects were estimated from linear mixed models with a pedigree-derived kinship matrix as the random-effect covariance and age and sex as fixed effects. The polygenic variance component approached zero in all four outcomes, indicating that carrier status itself accounted for the family-clustered variance; inference is reported with t statistics on n − 4 degrees of freedom. Abbreviations: CI = confidence interval; D = diopter; miR-184 = microRNA-184; SD = standard deviation.

| Outcome | Affected | Unaffected | Carrier effect<br>(95% CI)* | P value |
| --- | --- | --- | --- | --- |
| | mean ± SD | mean ± SD | | |
| Axial length<br>(mm) | 22.26 ± 1.24 | 23.29 ± 0.57 | –2.22 (–4.03, –0.40) | 0.022 |
| Mean<br>keratometry (D) | 52.16 ± 3.45 | 43.91 ± 0.69 | +9.30 (+3.83, +14.78) | 0.004 |
| White-to-white<br>(mm) | 10.43 ± 0.28 | 12.02 ± 0.16 | –1.61 (–2.11, –1.10) | <0.001 |
| Central<br>pachymetry (μm) | 414.9 ± 30.2 | 539.0 ± 24.4 | –138.7 (–210.6,<br>–66.8) | 0.003 |

The polygenic variance component approached zero in all four outcomes, indicating that carrier status itself accounted for the family-clustered variance in these measurements; a sensitivity analysis restricted to one individual per sibship (n = 6–7) yielded effect estimates in the same direction and of similar magnitude, further confirming the findings: axial length −1.65 mm (95% CI −5.49 to +2.19; p = 0.27), mean keratometry +11.52 D (+7.42 to +15.63; p = 0.003), white-to-white −1.41 mm (−2.80 to −0.02; p = 0.048), and central pachymetry −111.7 µm (−328.2 to +104.8; p = 0.16). Widening confidence intervals reflect the reduced sample size.

### Cellular and Molecular Phenotype

To substantiate the use of corneal endothelium as the cellular substrate for mechanistic studies of EDICT syndrome, we first compared miR-184 expression between primary corneal endothelial and scleral tissue from a single donor (Supplemental Figure S2A). The DMEK graft demonstrated approximately 2500-fold higher miR-184 expression than undifferentiated iPSCs from a control sample (mean normalized 2^−ΔΔCt^ = 2483). The scleral rim also showed detectable expression of miR-184 but approximately 5.5-fold lower expression relative to corneal endothelium. The quantitative TaqMan assay also confirmed that CECs differentiated in vitro from iPSCs upregulate miR-184 expression ∼200-fold relative to pre-differentiation iPSC. The specificity of the TaqMan detection assay as well as the significant upregulation of miR-184 upon CEC differentiation were confirmed by northern blot analysis (Supplemental Figure S2B).

To assess cellular and molecular pathogenesis, we generated one patient-derived iPSC line and differentiated it alongside three control iPSC lines (JHiPS-30516, JHiPS-70226, JHiPS-70351) into CECs across three independent differentiation batches per line. Immunostaining for ZO-1 in mutant CECs revealed irregular, discontinuous cell borders and defective intercellular junctions (Figure 2A). Quantitative analysis confirmed increased cell area (Figure 2C), larger nucleus area (Figure 2D), and greater intercellular gaps (Figures 2E, 2F). Staining for ATP1A1 identified abnormal membrane architecture (Figure 2B), which appeared patchy and disrupted; metrics showed an increase in patch number (Figure 2G), patch density (Figure 2H), peak number (Figure 2I), and decrease in peak intensity (Figure 2J), reflecting abnormal membrane protein distribution and possibly impaired pump function.

**Figure 2.**
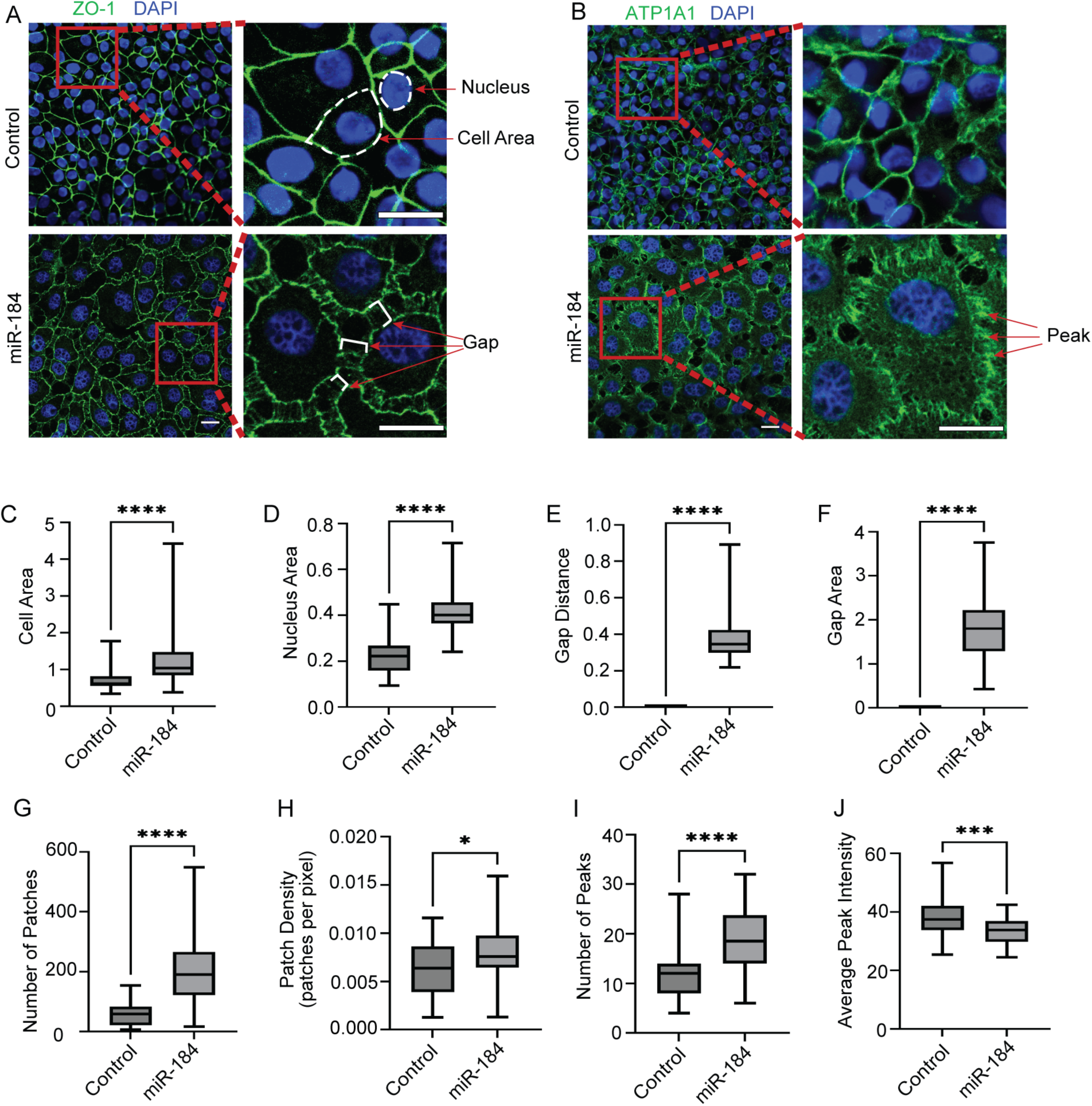
Immunofluorescent staining and quantitative analysis of control and miR-184 mutant iPSC-derived CEC. **(A, C-F)** Immunofluorescent staining and quantitative analysis of control and miR-184 mutant iPSC-derived CECs for ZO-1 and DAPI. Scale bars: 10 μm. Box-and-whisker plot showing the median of the relative cell area(C) of control hiPSC-derived CEC (n =60 cells from three replicates) compared to miR-184 mutant hiPSC-derived CEC (n =50 cells from three replicates), the relative nucleus area(D) of control hiPSC-derived CEC (n =72 cells from three replicates) compared to miR-184 mutant hiPSC-derived CEC (n =50 cells from three replicates), the relative gap distance(E) of control hiPSC-derived CEC (n =30 cells from three replicates) compared to miR-184 mutant hiPSC-derived CEC (n =52 cells from three replicates), the relative gap area(F) of control hiPSC-derived CEC (n =30 cells from three replicates) compared to miR-184 mutant hiPSC-derived CEC (n =49 cells from three replicates). Error bars represent the interquartile range. p value was calculated using a two-tailed unpaired t test. **(B, G-J)** Immunofluorescent staining and quantitative analysis of control and miR-184 mutant iPSC-derived CECs for ATP1A1 and DAPI. Note the patchiness of mutant cells and an increase in gaps. Scale bars: 10 μm. Box-and-whisker plots showing the median of the relative number of patches(G), the relative patch density(H), the relative number of peaks(I) and the relative average peak intensity(J) of control hiPSC-derived CEC (n =45 cells from three replicates) compared to miR-184 mutant hiPSC-derived CEC (n =50 cells from three replicates). Error bars represent the interquartile range. p value was calculated using a two-tailed unpaired t test. Abbreviations: ATP1A1 = adenosine triphosphatase Na+/K+ transporting subunit alpha 1; CEC = corneal endothelial cell; DAPI = 4′,6-diamidino-2-phenylindole; iPSC = induced pluripotent stem cell; ZO-1 = zonula occludens-1.

Gene expression profiling in mutant CECs (Figures 3A–I) demonstrated significant downregulation of COL4A1, COL4A3, AQP1, ATP1A1, FOXC1, and SLC16A3 and upregulation of COL8A1, molecules essential for Descemet’s membrane integrity, endothelial cell adhesion, and fluid regulation, and commonly implicated in endothelial dystrophies.

**Figure 3.**
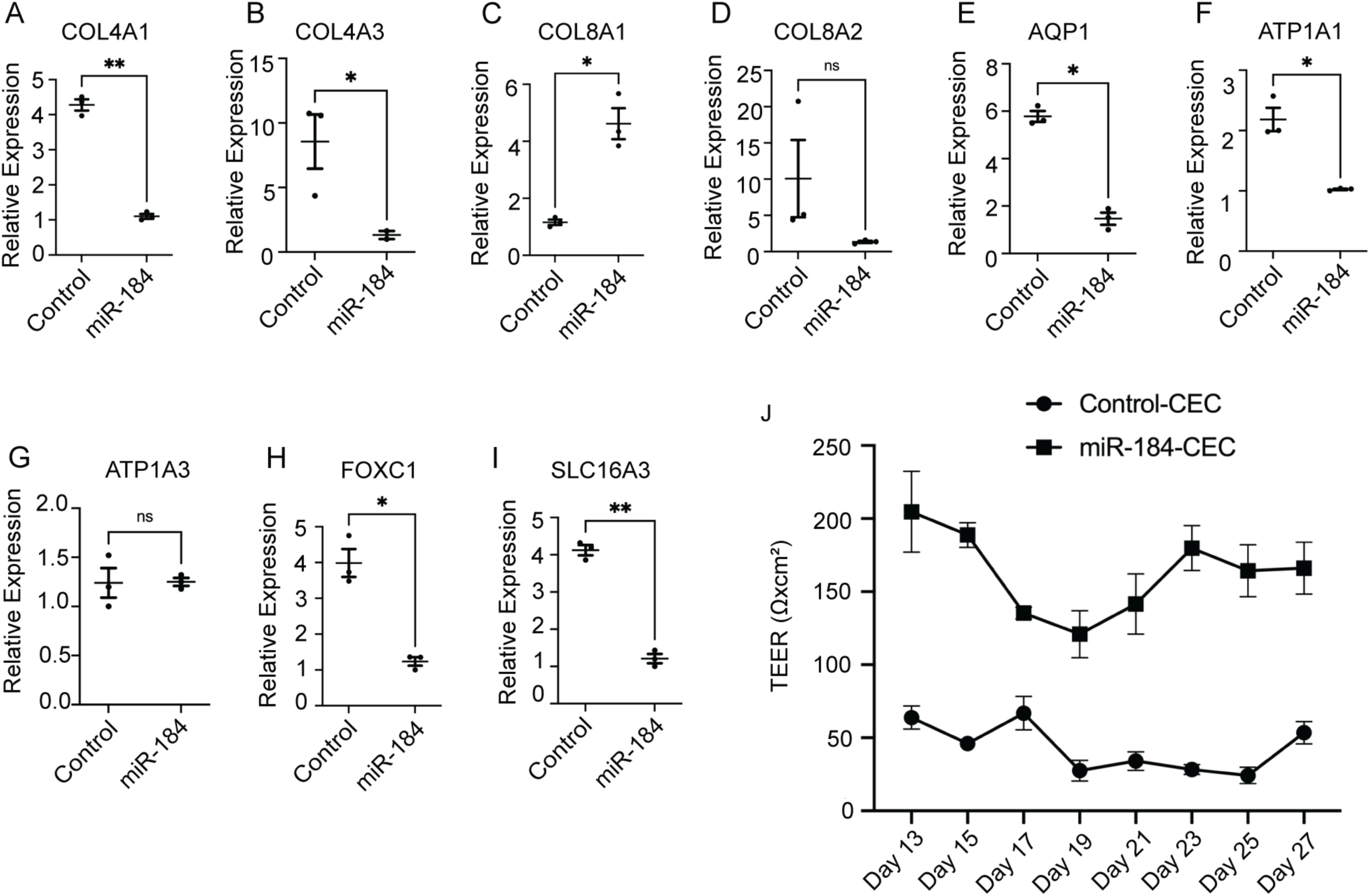
The miR-184 variant is associated with altered function of iPSC-derived CECs. (A–I) Relative expression of COL4A1, COL4A3, COL8A1, COL8A2, AQP1, ATP1A1, ATP1A3, FOXC1, and SLC16A3 in control and miR-184 mutant iPSC-derived CECs, as measured by qRT-PCR. Error bars show SEM; statistical significance is shown for each comparison (n = 3 independent differentiation batches). P values were calculated using two-tailed unpaired t tests. *P < 0.05; **P < 0.01; ns = nonsignificant. (J) TEER assay values measured from days 13 to 27 during CEC differentiation and maturation. Error bars show mean ± SD (n = 3 independent differentiation batches). Abbreviations: AQP1 = aquaporin 1; ATP1A1 = adenosine triphosphatase Na+/K+ transporting subunit alpha 1; ATP1A3 = adenosine triphosphatase Na+/K+ transporting subunit alpha 3; CEC = corneal endothelial cell; COL4A1 = collagen type IV alpha 1 chain; COL4A3 = collagen type IV alpha 3 chain; COL8A1 = collagen type VIII alpha 1 chain; COL8A2 = collagen type VIII alpha 2 chain; EDICT = endothelial dystrophy, iris hypoplasia, congenital cataract, and stromal thinning; FOXC1 = forkhead box C1; iPSC = induced pluripotent stem cell; miR-184 = microRNA-184; ns = nonsignificant; qRT-PCR = quantitative reverse-transcription polymerase chain reaction; SD = standard deviation; SEM = standard error of the mean; SLC16A3 = solute carrier family 16 member 3; TEER = transendothelial electrical resistance.

Functional assessment of barrier function via TEER showed decreased resistance throughout days 13 to 30 of differentiation (Figure 3J), paralleling loss of junctional integrity and consistent with disrupted corneal endothelial monolayer functionality. These findings, confirmed in two additional healthy control iPSC lines (Supplemental Figure S3), suggest that the miR-184 variant is associated with altered CEC morphology and gene expression, accompanied by reduced endothelial barrier function.

## Discussion

In this study, we extend the clinical spectrum associated with the +57C>T substitution in the seed region of miR-184 beyond the originally described anterior segment dysgenesis to include microphthalmia, retinal detachment, and more generalized disruption of ocular development. Notably, the unique combination of a higher risk of retinal detachment and reduction in axial length indicates that this variant exerts effects on both anterior and posterior segment morphogenesis.

The coexistence of increased retinal detachment and shorter axial length may seem counterintuitive, as rhegmatogenous detachment is frequently associated with myopic axial elongation. A plausible explanation is that the variant produces primary developmental abnormalities of the retina or vitreoretinal interface that predispose to retinal breaks independent of axial myopia. Future high-resolution retinal imaging and histologic studies will define the posterior segment phenotype more precisely.

Ocular biometry in affected individuals demonstrated a significantly shorter axial length with a mean difference of 2.2 mm compared with unaffected relatives after adjusting for age, sex, and familial relationships. While axial length is generally multifactorial, the magnitude of the difference cannot be explained by corneal flattening alone. Together with the reduced horizontal corneal diameter and decreased central and peripheral corneal thickness, these data indicate a phenotype of overall smaller eyes. This is concordant with findings from miR-184 knockout zebrafish, which develop microphthalmia and lens opacification, further supporting a conserved role for miR-184 in regulating globe size and anterior segment architecture.^6^

A question raised by earlier reports of miR-184 variants is whether the observed corneal steepening represents a form of corneal thinning or keratoconus. In one family, the proband was reported to have keratoconus, but changes appeared in early childhood and no topography images were published.^4,11^ In our cohort, Scheimpflug imaging showed a regular bow-tie astigmatic pattern starting in childhood without inferior focal steepening, and the thinning extended beyond the central cornea to the periphery. These features are consistent with a primary corneal thinning disorder rather than keratoconus. One possible explanation for why keratoconus appeared in the proband in another family^5^ is that the diffuse corneal thinning could increase susceptibility to other keratoconus-associated genetic mutations that would otherwise be masked in a thicker cornea. Overall, these findings highlight a phenotype distinct from keratoconus. They point to the need for well-characterized clinical data with adequate statistical power, and the importance of consistent diagnostic criteria for keratoconus.

miR-184 is one of the most highly conserved ocular microRNAs and is expressed predominantly in the corneal and lens epithelia, where it contributes to the specification and maintenance of avascular, transparent ocular surfaces.^4,12,13^ Notably, affected individuals in this family did not demonstrate increased corneal neovascularization, suggesting that the +57C>T variant does not simply result in loss of function or frank haploinsufficiency. Future work quantifying relative wild-type and mutant miR-184 abundance using the TaqMan miRNA assay will help delineate these mechanisms.

In this study, we focused our experiments on the corneal endothelium. Relative to sclera, we found a cornea-predominant expression pattern consistent with prior reports describing miR-184 as enriched in ocular surface tissues^4,12,13^ Some affected individuals in this cohort had been initially diagnosed with Fuchs dystrophy, given the clinical appearance of a corneal endothelial dystrophy with guttae. In fact, a recent study identified alternative mutations in miR-184, the n.58G>A and n.73G>T variants, in a large cohort of patients with patients diagnosed with Fuchs dystrophy^6^ but who did not harbor the trinucleotide repeat expansion typically associated with the disease.^14^ That study reported widespread gene-expression perturbations consistent with endothelial dysfunction. In contrast to transcriptome-only profiling, our data provide functional corroboration at the cellular level, including disrupted junctional organization and reduced barrier function (TEER), together supporting a model in which miR-184 seed-region disruption compromises endothelial integrity. Taken together, the transcriptomic and functional phenotypes converge on a corneal endothelial mechanism while motivating future studies in posterior-segment–relevant models to address retinal detachment risk.

One limitation of this study is that mechanistic studies were performed using a single patient-derived line and multiple control lines; however, the magnitude of difference seen between affected and unaffected samples was profound and unexplained by normal variability. Future studies using isogenic correction or additional affected family members will be necessary to confirm these findings.

While our mechanistic studies were performed in iPSC-derived CECs, which enables rigorous testing of endothelial junctional integrity, extracellular matrix–related gene expression, and barrier function, a limitation is that it does not directly interrogate retinal cell types or vitreoretinal interface biology. Accordingly, the association between +57C>T miR-184 and retinal detachment in this pedigree should be interpreted as a clinical expansion of phenotype rather than proof of a retinal cell-autonomous mechanism derived from the CEC model. Future studies will require retinal and/or vitreoretinal interface–relevant iPSC-derived models (e.g., retinal pigment epithelium, retinal organoids, or co-culture systems) to define the posterior segment mechanism(s).

Taken together, the human and experimental data support a framework in which heterozygosity for the miR-184 +57C>T variant is associated with disrupted corneal endothelial junctional integrity and altered extracellular matrix-related gene expression, with a pan-ocular phenotype encompassing microphthalmia, corneal thinning, and susceptibility to retinal detachment. These findings highlight the centrality of miR-184 to coordinated ocular development and underscore the need for continued mechanistic studies aimed at identifying targets for future therapeutic modulation.

## Data Availability Statement

The data supporting the findings of this study are available from the corresponding author upon reasonable request. Individual-level clinical and genetic data are not publicly available because of privacy considerations.

## Abbreviations/Acronyms

microRNA: micro-ribonucleic acid
miR-184: microRNA-184
EDICT: endothelial dystrophy, iris hypoplasia, congenital cataract, and stromal thinning
iPSC: induced pluripotent stem cell
CEC: corneal endothelial cell
ZO-1: zonula occludens-1
ATP1A1: adenosine triphosphatase Na+/K+ transporting subunit alpha 1
qRT-PCR: quantitative reverse-transcription polymerase chain reaction
TEER: transendothelial electrical resistance
COL4A1: collagen type IV alpha 1 chain
COL4A3: collagen type IV alpha 3 chain
AQP1: aquaporin 1
COL8A1: collagen type VIII alpha 1 chain
OMIM: Online Mendelian Inheritance in Man
IRB: Institutional Review Board
JHUSOM: Johns Hopkins University School of Medicine
PBMC: peripheral blood mononuclear cell
ISCRO: Institutional Stem Cell Research Oversight
DMEK: Descemet membrane endothelial keratoplasty
RNU6A: RNA, U6 small nuclear 1
U6: U6 small nuclear RNA
snRNA: small nuclear RNA
ssDNA: single-stranded DNA
EDTA: ethylenediaminetetraacetic acid
SDS: sodium dodecyl sulfate
SSC: saline-sodium citrate
DMEM-F12: Dulbecco modified Eagle medium/F12
KSR: knockout serum replacement
bFGF: basic fibroblast growth factor
PDGF-BB: platelet-derived growth factor-BB
DKK-2: Dickkopf-related protein 2
PBS: phosphate-buffered saline
PBST: phosphate-buffered saline with Triton X-100
DAPI: 4′,6-diamidino-2-phenylindole
GAPDH: glyceraldehyde-3-phosphate dehydrogenase
SEM: standard error of the mean
SD: standard deviation
COL8A2: collagen type VIII alpha 2 chain
ATP1A3: adenosine triphosphatase Na+/K+ transporting subunit alpha 3
FOXC1: forkhead box C1
SLC16A3: solute carrier family 16 member 3.

## Notes

This work was supported by the Maryland Stem Cell Research Fund (JX, AE), the Anderson Family Fund (AE), the Tolsma Family Fund (AE), the Franks Family Fund (AE), the Johns Hopkins Alzheimer’s Disease Research Center Faculty Award, NIH grant AG066507 (JX), and an unrestricted grant from Research to Prevent Blindness (AE). The sponsor or funding organization had no role in the design or conduct of this research.

### Competing Interest Statement

Y. Huang, M. Sulewski, J. Xu, and A.O. Eghrari report an ownership interest in Revivify Innovations. All other authors declare no competing interests. The funding organizations had no role in the design or conduct of the research.

### Author Declarations

The Institutional Review Board of the Johns Hopkins University School of Medicine gave ethical approval for this work. Written informed consent was obtained from all participants prior to enrollment. All experiments involving human induced pluripotent stem cells were conducted under oversight by the Johns Hopkins University Institutional Stem Cell Research Oversight Committee.

