## Supplemental Figure S1 for "The +57C>T substitution in microRNA-184 is associated with microphthalmia, retinal detachment, and altered ocular development"

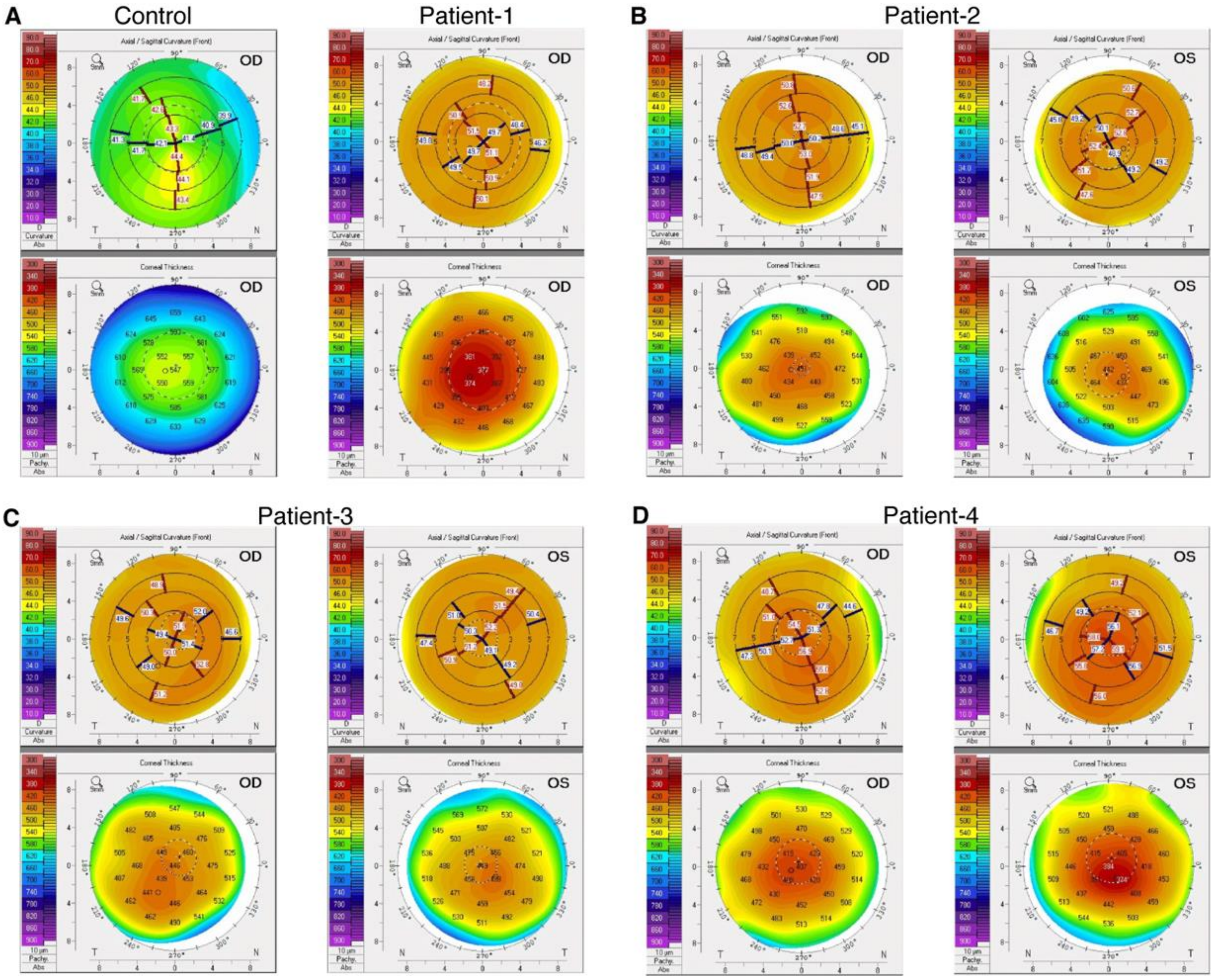


***Supplementary Figure S1.*** *Additional partial coherence inferometry and corneal topography with Scheimpflug imaging. (A) Corneal topography of control eyes from unaffected family members using Scheimpflug imaging. (B-D) Corneal topography of control eyes from affected family members using Scheimpflug imaging. Corneas are diffusely thin in both the center and periphery and with smaller diameter, unlike focal, apical thinning seen in classic keratoconus. Abbreviations: PCI = partial coherence interferometry*
