## Supplemental Figure S2 for "The +57C>T substitution in microRNA-184 is associated with microphthalmia, retinal detachment, and altered ocular development"

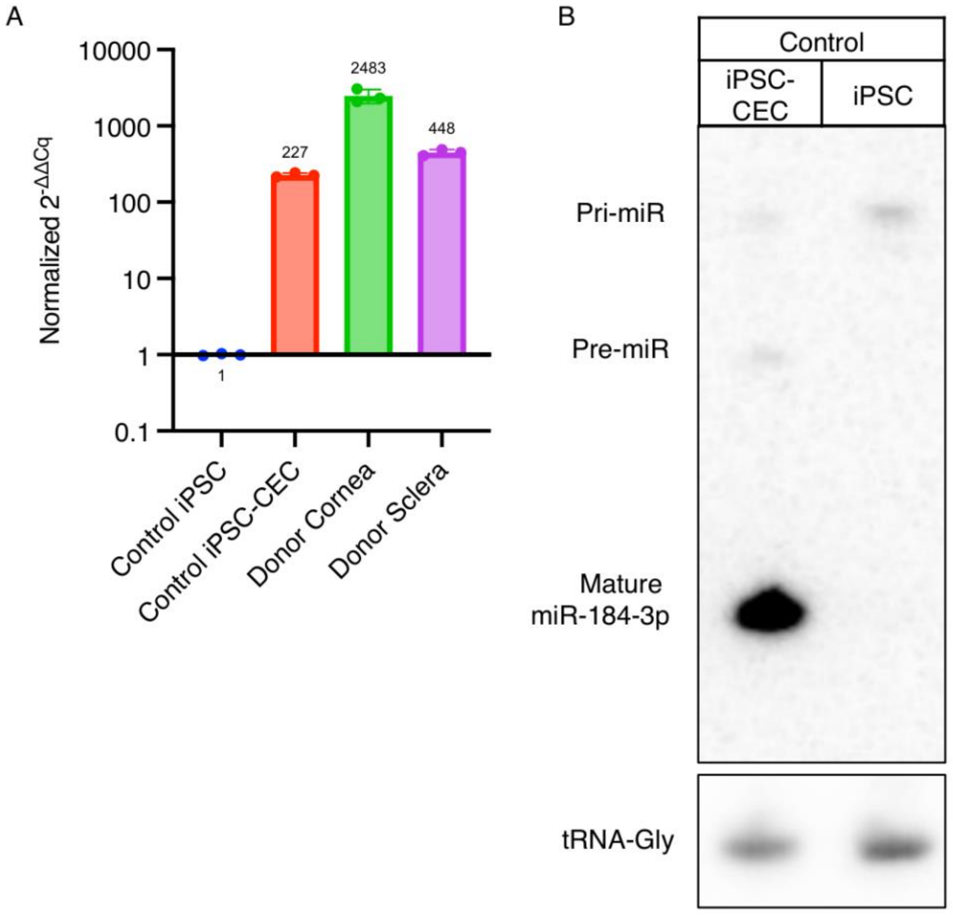


***Supplemental Figure S2.*** *(A) TaqMan MicroRNA quantification assay for mature miR-184-3p. Every sample was normalized to the abundance of U6 RNA quantified by TaqMan assay. The normalized relative abundance of miR-184 was calculated relative to non-differentiated iPSCs, which express background level of miR-184. iPSC-derived CECs (14 days post-differentiation) upregulate miR-184 expression ~200-fold relative to pre-differentiation iPSCs. miR-184 was also measured in the cornea and sclera of an individual donor, showing that miR-184 is ~5-fold higher in corneal endothelium than in the corresponding sclera. n=3 technical replicates per sample, bars represent mean and SD. (B) Northern blot analysis of total RNA from iPSCs and iPSC-derived CECs (14 days post-differentiation), from a control sample. Radiolabeled probe against mature miR-184 also detects partially processed pre-miR-184 and primary transcript. A probe against tRNA-Gly was used as a loading control. Abbreviations: CEC = corneal endothelial cell; DMEK = Descemet membrane endothelial keratoplasty; iPSC = induced pluripotent stem cell; microRNA-184 = micro-ribonucleic acid 184; tRNA-Gly = transfer ribonucleic acid-glycine; pri-miR = primary micro-ribonucleic acid; pre-miR = precursor micro-ribonucleic acid*
