## Supplemental Figure S3 for "The +57C>T substitution in microRNA-184 is associated with microphthalmia, retinal detachment, and altered ocular development"

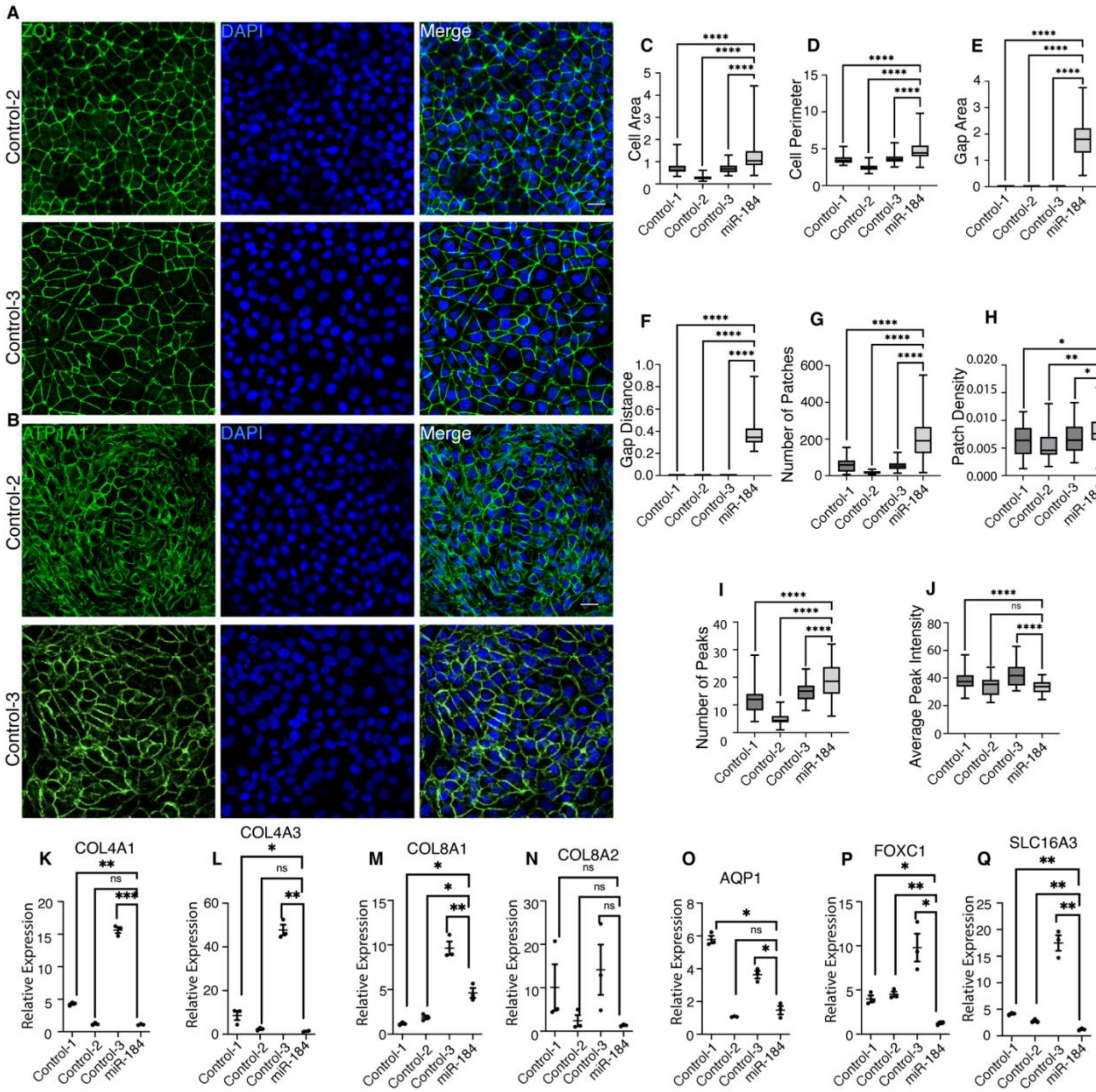


**Supplementary Figure S3. Additional immunofluorescent staining/qPCR and quantitative analysis of control and mir-184 mutant iPSC-derived CECs** (A and B) Fluorescent imaging of ZO1 and ATP1A1 of iPSC-derived CECs created from two additional healthy control donors. Scale bars: 10 µm. (C-J) Quantitative analysis of iPSC-derived CECs showing characterization of irregular mutant phenotype as compared to cells from three healthy controls. (K-Q) Changes in relative qPCR expression of mutant vs three controls for seven CEC related genes. Error bars indicate SEM. Statistical comparisons were performed by 2-way ANOVA for multiple comparisons. $^{*}P < 0.05$, $^{**}P < 0.01$, $^{***}P < 0.001$, $^{****}P < 0.0001$. Abbreviations: AQP1 = aquaporin 1; ATP1A1 = adenosine triphosphatase Na+/K+ transporting subunit alpha 1; CEC = corneal endothelial cell; COL4A1 = collagen type IV alpha 1 chain; COL4A3 = collagen type IV alpha 3 chain; COL8A1 = collagen type VIII alpha 1 chain; COL8A2 = collagen type VIII alpha 2 chain; DAPI = 4′,6-diamidino-2-phenylindole; FOXC1 = forkhead box C1; iPSC = induced pluripotent stem cell; ns = nonsignificant; SLC16A3 = solute carrier family 16 member 3; ZO-1 = zonula occludens-1.
